# The Proportion of Randomized Controlled Trials That Inform Clinical Practice: A Longitudinal Cohort Study of Trials Registered on ClinicalTrials.gov

**DOI:** 10.1101/2022.05.12.22275021

**Authors:** Nora Hutchinson, Hannah Moyer, Deborah A. Zarin, Jonathan Kimmelman

## Abstract

**Background:** Prior studies suggest that clinical trials are often hampered by problems in design, conduct and reporting that limit their uptake in clinical practice. We have described “informativeness” as the ability of a trial to guide clinical, policy or research decisions. Little is known about the proportion of initiated trials that inform clinical practice.

**Methods:** We created a cohort of randomized interventional clinical trials in three disease areas (ischemic heart disease, diabetes mellitus and lung cancer), that were initiated between 1 January 2009 and 31 December 2010 using ClinicalTrials.gov. We restricted inclusion to trials aimed at answering a clinical question related to the treatment or prevention of disease. Our primary outcome was the proportion of clinical trials fulfilling four conditions of informativeness: importance of the clinical question, trial design, feasibility, and reporting of results.

**Results:** Our study included 125 clinical trials. The proportion meeting four conditions for informativeness was 26.4% (95% CI 18.9 – 35.0). Sixty-seven percent of participants were enrolled in informative trials. The proportion of informative trials did not differ significantly between our three disease areas.

**Conclusions:** Our results suggest that the majority of clinical trials designed to guide clinical practice possess features that may compromise their ability to do so. This highlights opportunities to improve the scientific vetting of clinical research.

**Funding:** This study was funded by the Fonds de recherche Santé Québec postdoctoral research grant (NH). This funding body was not involved in study design, conduct or reporting.

## Introduction

The ultimate goal of clinical research is to produce evidence that supports clinical and policy decisions. Numerous analyses suggest that a substantial proportion of clinical trials aimed at informing clinical practice are marred by flaws in design, execution, analysis and reporting.^1–8^ The initial research response to COVID-19 illustrated the fact that existing oversight mechanisms fail to prevent the initiation of flawed trials.^9^ While unexpected events can stymie well-conceived and implemented studies, trials that have features rendering them unlikely to inform clinical practice may do harm by misleading potential participants of their benefits, and by diverting patient-participants from otherwise informative research efforts.^10^

We have previously described five conditions that trials should fulfill to support clinical or policy decision-making.^10,11^ First, trials must ask an important and clinically relevant question that is not yet resolved. Second, trials must be designed to provide a meaningful answer to that question. Third, trials must be feasible, with achievable enrollment goals and timely primary outcome completion. Fourth, outcomes must be analyzed in ways that support valid interpretation. Last, trial results must be made accessible in a timely fashion.

In what follows, we created surrogate measures for four conditions of informativeness: trial importance, design quality, feasibility, and reporting (the fifth condition, analytical integrity, did not lend itself to objective, dichotomous assessment, and is not assessed below). We then evaluated the proportion of “clinically directed randomized controlled trials” in three common disease areas meeting these four conditions. This information can be used to help healthcare and research systems identify studies in need of further scrutiny, thereby improving the impact of their research.

## Methods

### Overview of Approach

We created a cohort of randomized, interventional clinical trials in three broad disease areas that are representative of the clinical research enterprise and that have a significant impact on patient morbidity and mortality: ischemic heart disease, diabetes mellitus and lung cancer. We restricted inclusion to trials that appeared to be aimed at informing clinical practice by selecting trials with a stated purpose of treatment or prevention of disease and with a primary clinical outcome or appropriate surrogate. We established milestones that could serve as objectively verifiable surrogates for four conditions of informativeness. Trials in our sample were then tracked forward to assess the proportion attaining each informativeness condition. “Informative trials” were trials that fulfilled all four conditions of informativeness.

### Surrogate Measures for Four Conditions of Informativeness

We formulated surrogate measures for each condition of informativeness. Measures were chosen based on i) close correspondence with each informativeness condition; ii) objective and reproducible dichotomous scoring; and iii) feasibility of assessment. The four surrogates of informativeness, described in greater detail below, were as follows: trial importance (determined by citation of reported trial results in high quality clinical synthesizing documents; the premise of this surrogate is that these documents focus on questions of clinical importance); trial design quality (assessed using a modified Cochrane risk of bias (ROB) tool which is designed to identify threats to study internal validity); trial feasibility (established based on ability to achieve adequate participant enrollment and timely primary outcome completion); and reporting (based on accessibility of primary outcome results via deposition on ClinicalTrials.gov or in journal publications).

### Clinical Trial Sampling

We identified all trials registered on ClinicalTrials.gov in our three disease areas with a start date from 1 January 2009 to 31 December 2010 inclusive (eMethods 1 – search criteria). Our time range provided a minimum of nine years of follow-up for maturation toward trial completion and fulfillment of all four surrogates of informativeness. Trials were downloaded from ClinicalTrials.gov on 15 May 2020. We updated trial status and enrollment for all trials meeting our inclusion criteria on 6 October 2021.

We included randomized trials i) evaluating interventions of any type; ii) aimed at the treatment or prevention of ischemic heart disease, diabetes mellitus or lung cancer; iii) with at least one site in the United States (most of which will thus have a regulatory requirement for results reporting);^12^ and iv) interventions that were FDA approved, that advanced to FDA approval, or interventions not subject to FDA approval (e.g. cardiac rehabilitation). We did not include trials that we deemed unlikely to be targeted at informing clinical practice by excluding: i) studies that exclusively evaluated safety, diagnostic or screening interventions; and ii) early phase trials (phase 0 or phase 1) (eMethods 2 – inclusion/exclusion criteria; Figure 1 – flow diagram for trial inclusion; eMethods 3 – flow diagrams by disease; eMethods 4 – assessment of regulatory approval status). Phase 2 trials were included in our study as they are frequently used to inform both clinical and regulatory decision-making, particularly in cancer, where over one quarter of recent FDA cancer drug approvals were based on the results of phase 1/2 or phase 2 clinical trials.^13^ Trials were independently screened and assessed for eligibility by two authors (NH & HM), with disagreements resolved by a third reviewer (JK).

**Figure 1.**
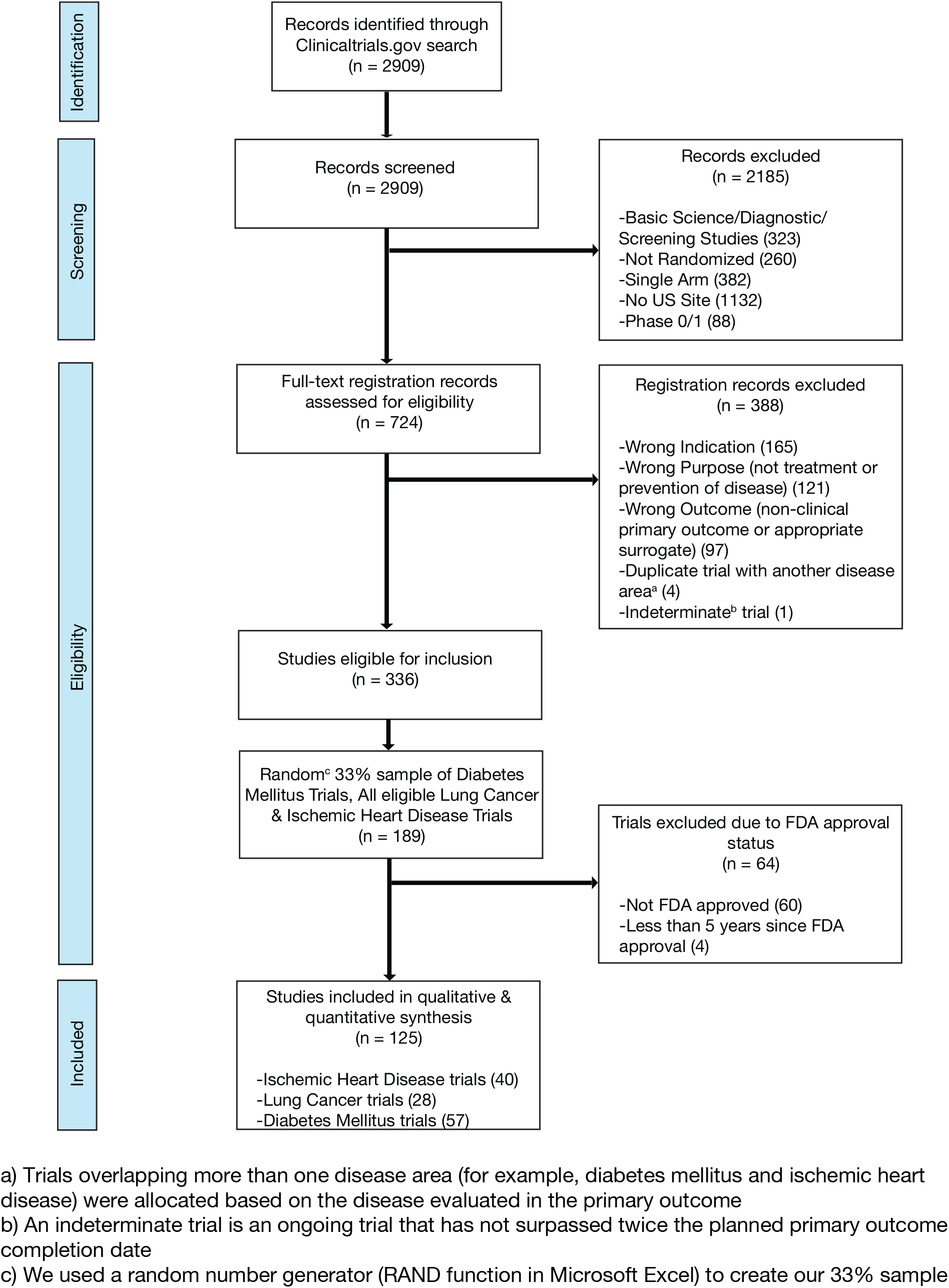
Flow Diagram for Trial Inclusion.

### Scoring Conditions of Informativeness

Two authors (NH & HM) independently scored all trials for the surrogate measures of the four conditions of informativeness (eTable 1; eMethods 5). Disagreements were resolved by a third reviewer (JK). Because of their logical relationship among surrogate measures (e.g. citation in a high quality clinical synthesizing documents cannot be assessed unless trial results are available) and workflow (e.g. risk of bias information is often available in systematic reviews), conditions were scored sequentially. Trials not meeting one condition were not advanced for evaluation of subsequent conditions. The order of scoring was as follows: i) feasibility; ii) reporting; iii) importance; and iv) design. Trials meeting all four conditions were deemed informative; trials failing on any condition possessed features that compromised their informativeness.

Our assessments of informativeness began by evaluating feasibility based on timely trial completion and patient-participant recruitment success. Terminated trials were deemed infeasible if the reason for termination in the ClinicalTrials.gov registration record involved accrual, feasibility, funding or another non-scientific reason (as opposed to termination due to accumulated scientific data suggesting early efficacy, futility or toxicity) (eMethods 6 – classification of reason for termination). Completed trials were deemed to have not fulfilled feasibility if final participant enrollment was less than 85% of expected enrollment as listed in the final registration record prior to study start, thus reflecting a substantial loss of statistical power for the primary outcome.^14^ Trials that were ongoing were categorized as infeasible if they had already surpassed double the intended time for primary completion, which was calculated by subtracting the intended primary completion date (as stated in the final registration record prior to study start) from the trial start date, then multiplying by two.

We next assessed results reporting by determining whether primary outcome results were publicly available. Trials were categorized as reported if they either had primary outcome results available on ClinicalTrials.gov or in a publication (eMethods 7 – methodology for publication search). When more than one publication presented primary outcome results, the earliest published report was identified and advanced to the next step of assessment. ClinicalTrials.gov results reporting and publication search were updated in October 2021 for those trials previously deemed to have not met the criteria for reporting.

Importance was scored by determining whether trial results were included in a high-quality review document designed to inform medical decision-making. To credit trials with being informative even if they produced negative results, trials were first assessed for inclusion in the results of a high-quality systematic review (SR), given that SR citation practices are results neutral. We assessed for trial results citation in a Cochrane SR, Agency for Healthcare Research and Quality (AHRQ) SR or in an SR deemed of high quality based on a modified AMSTAR score (eMethods 8 – SR search strategy and quality assessment). Trials not cited in the results of high-quality SRs were evaluated for inclusion in a high-quality CPG; remaining uncited trials were then assessed for inclusion in an UpToDate review article^15^ (eMethods 9 – CPG search strategy, CPG quality assessment and point-of-care medical database search). Trials cited in high-quality review documents were deemed to have fulfilled the importance condition. Assessment of importance was updated in October 2021 for all trials previously deemed to have not met the criterion for importance.

Finally, design was assessed by determining whether studies were at elevated risk of bias, using a modified Cochrane Risk of Bias (ROB) tool^16^ (eMethods 10). When available, ROB scores were extracted directly from high-quality SRs identified during the assessment of trial importance. When unavailable, ROB scores were independently performed by two authors (NH & HM), with disagreements resolved by a third reviewer (JK). Trials were deemed to have fulfilled the design condition of informativeness if all ROB elements were deemed to be of low risk of bias, or a majority were low risk of bias with a minority of elements deemed to be of unclear risk of bias.

### Statistical Analysis

Our primary outcome was the proportion of trials that met all four conditions of trial informativeness. We provided a 95% binomial confidence interval for our primary outcome. We performed a sensitivity analysis on our primary outcome excluding small, pilot-type studies that would not have been designed to inform clinical decision-making. These were identified based on an anticipated participant enrollment below the lowest quartile of target enrollment for our cohort of trials. Due to concern that phase 2 trials are less likely to inform clinical practice than trials of a higher phase, we performed a second sensitivity analysis on our primary outcome excluding phase 1/2 and phase 2 trials.

As secondary outcomes, we estimated the proportion of trial participants who were enrolled in informative trials, as well as the proportion of informative trials in each of our three disease areas. We also report the proportion of trials advancing across each condition of informativeness. We provided 95% binomial confidence intervals for these secondary outcomes.

We compared the proportion of informative trials between disease categories and by trial sponsor using the Chi-square test (chisq.test function in R) and provided binomial confidence intervals for each stratum. We used the fisher.test function in R to perform a two-sided Fisher’s Exact test assessing the proportion of informative trials by type of intervention and trial phase and provided exact confidence intervals for each. We calculated inter-rater agreement rates using Cohen’s kappa (eTable 2). We defined p < 0.05 as statistically significant. All analyses were performed using R version 4.0.2.^17^

Our study was not subject to Institutional Review Board approval, as it relied on publicly accessible data. The study protocol was prospectively registered on Open Science Framework;^18^ deviations and amendments to the study protocol are detailed in eMethods 11. The code^19^ and data set^18^ used in this analysis are available online. This study follows the Strengthening the Reporting of Observational Studies in Epidemiology (STROBE) reporting guidelines for cohort studies (eMethods 12).

## Results

Over half of the 125 interventional trials in our cohort were studies of drug or biologic interventions (77 trials; 61.6%). The majority were Phase 2 (24 trials, 19.2%) or Phase 3 trials (50 trials, 40.0%). Trial status was “Completed” in 99 of 125 trials (79.2%) and “Terminated” in 15 trials (12.0%) (Table 1).

**Table 1.** Characteristics of Intervention Trial Cohort.

| Category | Ischemic Heart Disease Trials<br>N = 40 | Diabetes Mellitus Trials<br>N = 57 | Lung Cancer Trials<br>N = 28 | All Trials<br>N = 125 (%) |
| --- | --- | --- | --- | --- |
| Trial Phase |  |  |  |  |
| 2<sup>a</sup> | 6 (15.0) | 5 (8.8) | 13 (46.4) | 24 (19.2) |
| 3<sup>b</sup> | 11 (27.5) | 26 (45.6) | 13 (46.4) | 50 (40.0) |
| 4 | 10 (25.0) | 9 (15.8) | 0 (0.0) | 19 (15.2) |
| NA | 13 (32.5) | 17 (29.8) | 2 (7.1) | 32 (25.6) |
| Intervention |  |  |  |  |
| Drug/Biologic | 19 (47.5) | 34 (59.6) | 24 (85.7) | 77 (61.6) |
| Combination<sup>c</sup> | 7 (17.5) | 0 (0.0) | 1 (3.6) | 8 (6.4) |
| Device | 4 (10.0) | 4 (7.0) | 0 (0.0) | 8 (6.4) |
| Other<sup>d</sup> | 10 (25.0) | 19 (33.3) | 3 (10.7) | 32 (25.6) |
| Trial Status |  |  |  |  |
| Completed | 29 (72.5) | 53 (93.0) | 17 (60.7) | 99 (79.2) |
| Terminated | 7 (17.5) | 1 (1.8) | 7 (25.0) | 15 (12.0) |
| Active, NR | 0 (0.0) | 1 (1.8) | 4 (14.3) | 5 (4.0) |
| Unknown | 4 (10.0) | 2 (3.5) | 0 (0.0) | 6 (4.8) |
| Outcome |  |  |  |  |
| Clinical | 24 (60.0) | 8 (14.0) | 10 (35.7) | 42 (33.6) |
| Surrogate | 16 (40.0) | 49 (86.0) | 18 (64.3) | 83 (66.4) |
| Sponsor<sup>e</sup> |  |  |  |  |
| Industry | 18 (45.0) | 27 (47.4) | 13 (46.4) | 58 (46.4) |
| Other | 22 (55.0) | 30 (52.6) | 15 (53.6) | 67 (53.6) |

<sup>a</sup>Including Phase 1/2
<sup>b</sup>Including Phase 2/3
<sup>c</sup>Including Drug + Device, Drug + Procedure, Behavioural + Device, Radiation Therapy + Drug
<sup>d</sup>Including Behavioural Intervention, Radiation Therapy, Surgical Procedure, Cellular Intervention
<sup>e</sup>As defined in ClinicalTrials.gov registration records

Our primary outcome, the proportion of trials that informed clinical practice, was 26.4% (95% CI 18.9 – 35.0) (Figure 2). As a sensitivity analysis, we re-analyzed our primary outcome excluding the 35 trials in the lowest quartile for target enrollment. This resulted in a proportion of informative trials of 35.6**%** (95% CI 25.7 – 46.3). We performed a second sensitivity analysis on our primary outcome excluding phase 1/2 and phase 2 trials. This resulted in 30.7% (95% CI 21.9 – 40.7) of trials meeting 4 conditions of informativeness.

**Figure 2.**
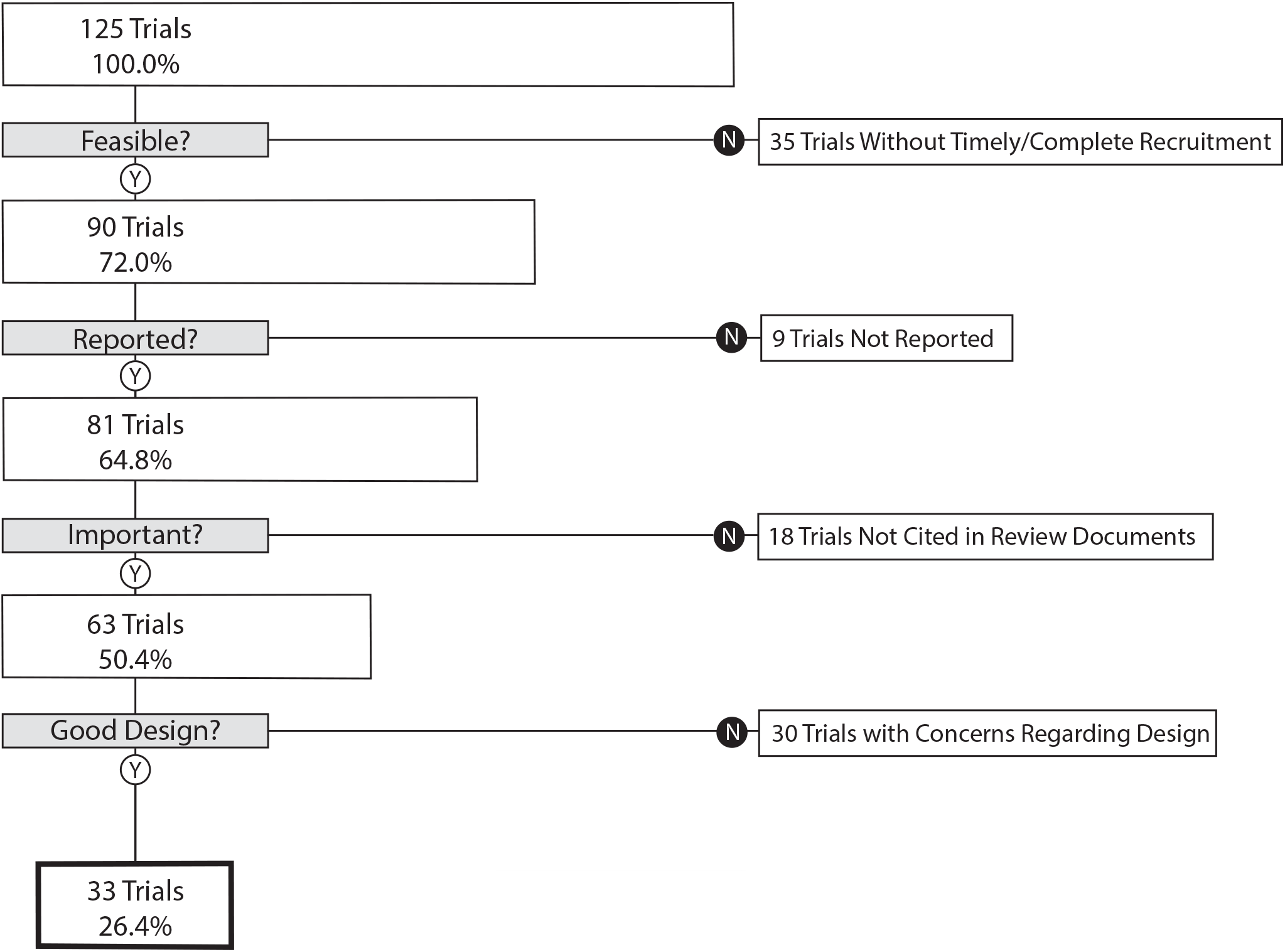
Flow Diagram - The Proportion of Trials Meeting Four Conditions of Informativeness.

A total of 193,839 participants were enrolled in the 125 trials in our cohort, of which 129,973 (67.1% (95% CI 66.8 – 67.3)) were enrolled in informative trials. The proportion of ischemic heart disease trials that was informative was 27.5% (95% CI 14.6 – 43.9); the proportion for diabetes mellitus trials was 31.6% (95% CI 19.9 – 45.2), and the proportion for lung cancer was 14.3% (95% CI 4.0 – 32.7) (Figure 3). Proportions did not vary significantly by disease area (p value = 0.23) (Figure 4). Each surrogate measure contributed considerably to the stepwise decline in the proportion of informative trials (eTable 3).

**Figure 3.**
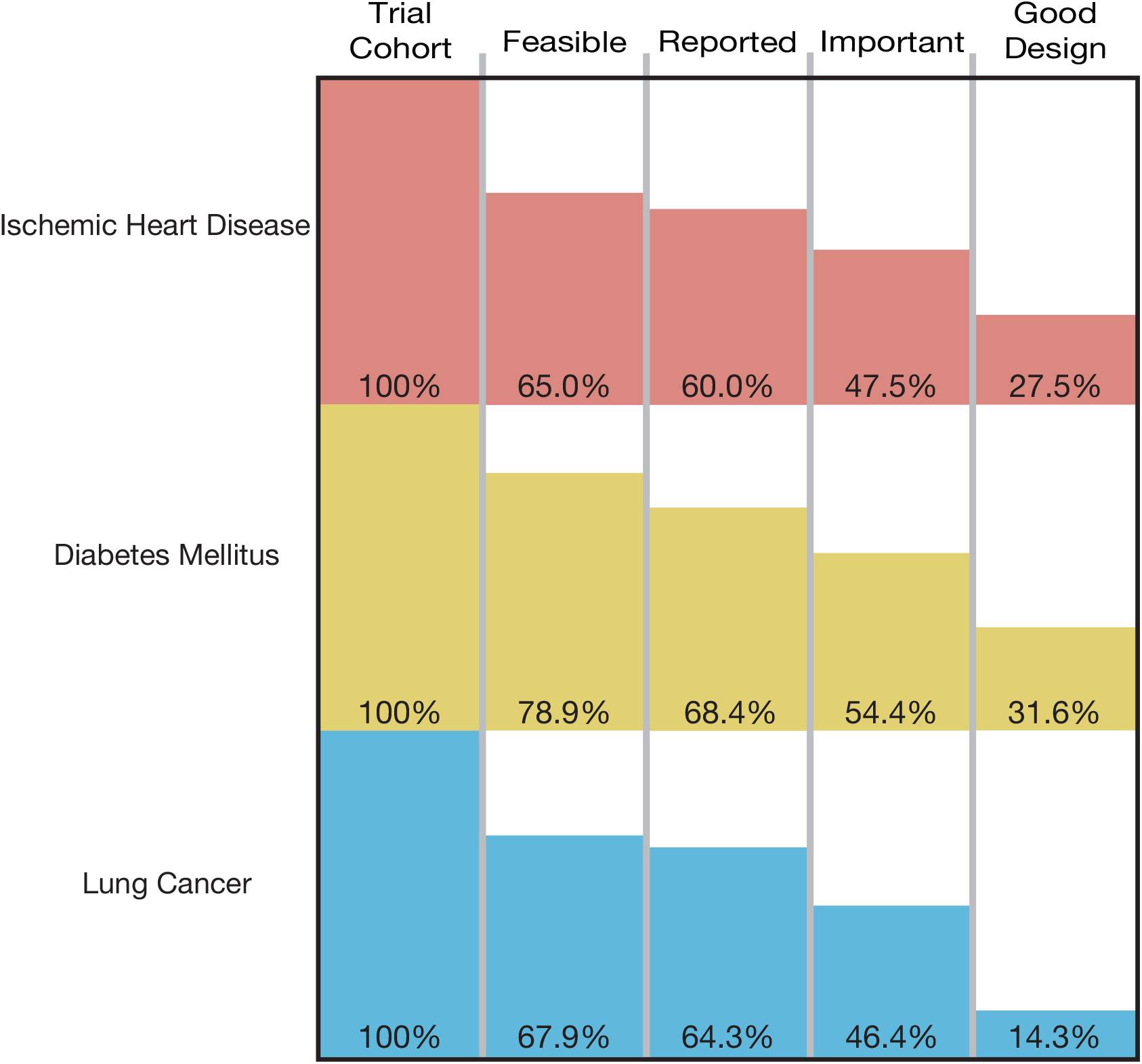
The Cumulative Proportion of Trials Meeting Four Conditions of Informativeness by Disease Area.

**Figure 4.**
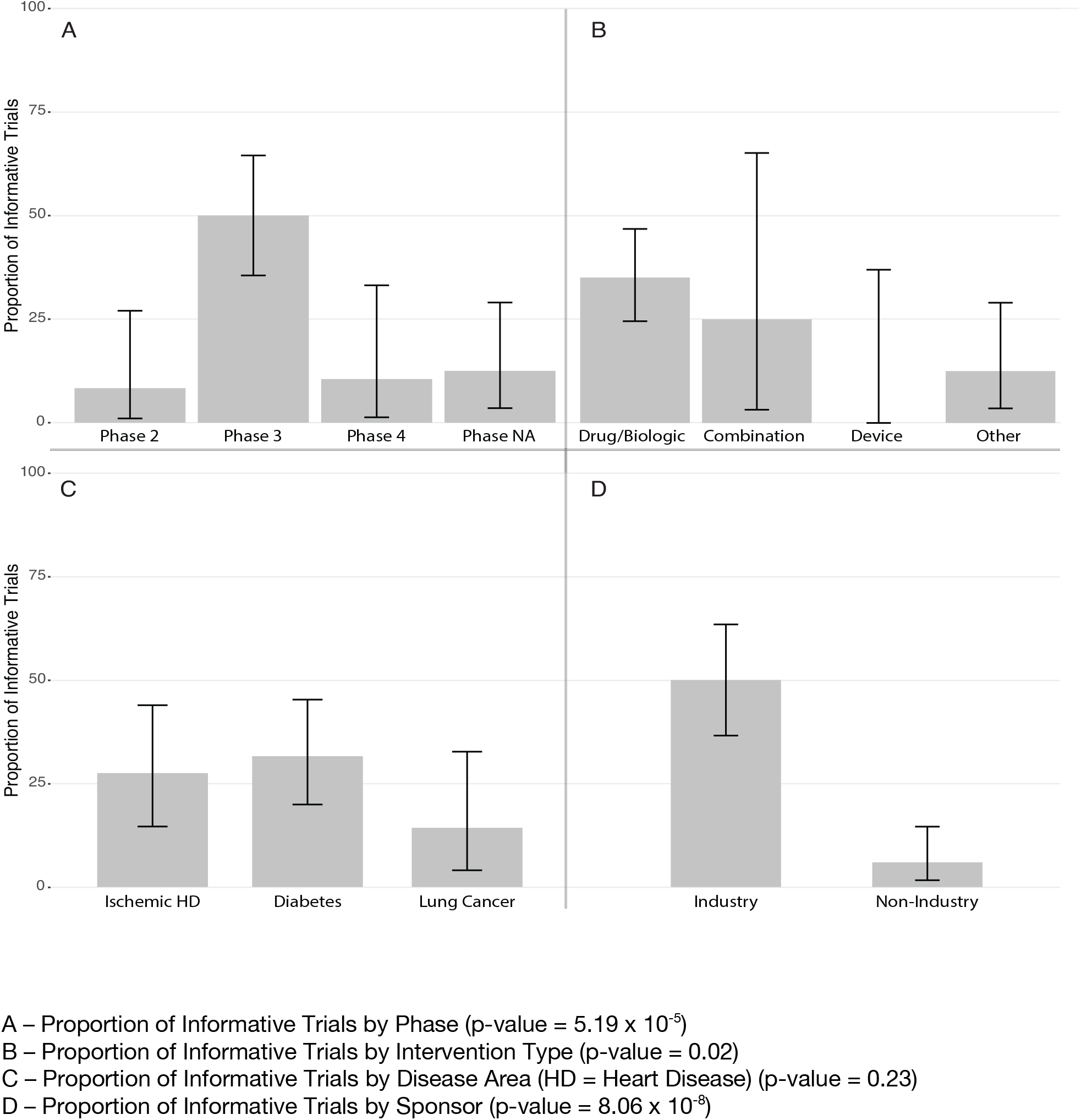
The Proportion of Informative Trials by Trial Property.

Studies sponsored by industry were significantly more likely to fulfill all four conditions of informativeness than those not sponsored by industry (50.0% vs. 6.0%, p value < 0.001)(Figure 4). Using the two-sided Fisher’s exact test, there was a non-random association between trial phase and informativeness, and type of intervention and informativeness (Figure 4).

## Discussion

This study provides the first assessment of the proportion of randomized trials fulfilling four key conditions of informativeness. In our analysis, just over one fourth of trials demonstrated adequacy for study feasibility, reporting, importance, and design. The remaining 73.6% contained a limitation in design, conduct or reporting that compromised their ability to inform clinical decision-making.

Certain shortcomings of clinical trials are a result of experimenting in a dynamic real-world environment and cannot be entirely avoided. Clinical trials are difficult to plan, and there may be defensible reasons for falling short of some conditions. For example, changes in medical practice may render a research question irrelevant to clinical practice; an emerging viral pandemic might lead to under-recruitment. However, our findings underscore the major challenges sponsors and clinical investigators confront in fulfilling the scientific and ethical warrant for enrolling patient-participants in randomized trials. The goal should be to address foreseeable limitations in trial design, conduct or reporting. For example, increased oversight by research funders, including requirements for landscape analysis of completed and ongoing clinical trials to ensure trials are addressing important questions, and the provision of independent scientific review to highlight vulnerabilities in trial design,^20^ are measures that can be implemented to increase the likelihood that trials will be informative. Many methodological weaknesses in trial design can be corrected at minor cost.^8^

The proportion of informative trials did not differ significantly between ischemic heart disease, diabetes mellitus and lung cancer, indicating shared challenges in design, implementation, and reporting. Our study also demonstrated that each condition of informativeness goes unfulfilled in roughly equal proportions (eTable 2), suggesting that vigilance is required throughout the life cycle of a trial. Our estimates for the fraction of studies fulfilling criteria for recruitment feasibility are in line with prior studies.^14,21–23^ The fraction of trials at low risk of bias is similar to prior estimates.^8,24,25^ Our estimate for the fraction of studies fulfilling reporting requirements (90.0%) is in line with prior studies that evaluated both ClinicalTrials.gov results deposition and publication,^26,27^ both of which were deemed acceptable means of results reporting in our study. To our knowledge, our study is the first to apply these conditions jointly to a sample of trials, in addition to assessing importance via citation in clinical synthesizing documents.

Our results also indicate that certain types of trials may be at greater risk for having their informativeness compromised. Phase 4 trials fared worse than Phase 3 trials, with only 2 of 19 fulfilling all 4 conditions of informativeness (eTable 4). Trials sponsored by industry funders were far more likely to fulfill all four conditions than those with non-industry sponsorship (50.0% vs. 6.0%). This is in keeping with prior research demonstrating greater recruitment challenges for non-industry funded trials,^14^ in addition to diminished compliance with timely results reporting on ClinicalTrials.gov.^28^

These results suggest that funding bodies and academic medical centers may not provide adequate resources for fulfilling the clinical mission of the trials they support. Several recent initiatives aim at improving various aspects of informativeness, including increased consideration given to the importance and clinical relevance of the research question, the evidentiary basis for proposed research, study registration and reporting, by many funders.^29^ The implementation of new frameworks, such as INQUIRE, developed to guide academic institutions in addressing waste in research, including assessments of research design, feasibility, transparency, relevance, and internal and external validity, if widely adopted, may lead to further improvements in research quality.^30^ The SARS-CoV-2 pandemic has highlighted both the susceptibility of our clinical research enterprise to substandard trials, while also showing what is possible with robust research vetting, coordination and collaboration.^31^

### Limitations

Our study should be interpreted considering several limitations. First, our measures for each condition of informativeness are proxies for the concepts they represent. For example, scoring trial importance required citation in a clinical synthesizing document. This measure may have erroneously classified some informative trials as at risk of being uninformative (e.g. trials that evaluate disease management in niche populations that are not addressed in practice guidelines or systematic reviews). It may also have misclassified some trials as informative (e.g. trials addressing already resolved clinical hypotheses, which might nevertheless be cited in systematic reviews). To the former, none of the 18 trials not fulfilling the importance condition involved niche populations (eTable 5). We also acknowledge that some trials may inform clinical practice despite failing our criteria. The DAPT Study (NCT00977938) was a large Phase 4 study that was deemed at high risk of bias in several high-quality systematic reviews.^32,33^ However, this study has had an important impact on the clinical management of antiplatelet therapy following drug-eluting stent placement.^34^ Our metrics are best understood as capturing factors that seriously (but not fatally) compromise a trial’s prospects of informing practice, and that are rectifiable. Second, we applied strict inclusion/exclusion criteria when identifying out cohort of “clinically directed randomized controlled trials,” thus limiting generalizability to other types of trials, including those involving diagnostics or interventions that do not advance to FDA approval. The latter would require different criteria, given their primary goal of informing regulatory decision-making. Third, we used a longitudinal approach, since conditions like publication or citation are only fulfilled after a study is completed. Changes in research practices or policy occurring over the last decade might produce different estimates for the proportion of randomized trials that are informative.

## Conclusions

Trial volunteers are generally told that their participation will advance clinical practice. However, one third (33%) of patient-participants in our study were enrolled in trials that possessed at least one feature that compromised their goal of informing clinical practice. Sponsors and investigators often face unforeseeable challenges, and trials with flaws in design and implementation occasionally uncover actionable insights. Nevertheless, research systems and oversight should address persistent barriers to fulfilling the societal mission of clinical research.

## Data Availability

Our manuscript relies on publicly available data from ClinicalTrials.gov. Our data set is available online on Open Science Framework (DOI 10.17605/OSF.IO/3EGKU) (reference 18).

https://osf.io/3egku/

## Supplementary Material

eMethods 1 – ClinicalTrials.gov search criteria

eMethods 2 – Trial Inclusion and Exclusion criteria

eMethods 3 – Flow Diagrams for Each Disease

eMethods 4 – Assessment of Regulatory Approval Status

eTable 1 – Addressing 4 Conditions for Informative Clinical Trials

eMethods 5 – Flow Chart of Informative Criteria Assessment

eMethods 6 – Classification of Reason for Termination

eMethods 7 – Methodology for Publication Search

eMethods 8 – Systematic Review Citation Search Strategy and Quality Assessment

eMethods 9 – Clinical Practice Guideline and Point-of-Care Medical Database Search Strategies and Quality Assessment

eMethods 10 – Operationalization of modified Cochrane Risk of Bias score

eMethods 11 – Deviations to the Study Protocol

eMethods 12 – STROBE Checklist for Cohort Studies

eTable 2 – Inter-rater Agreement Rates

eTable 3 – Proportion of Trials Meeting Each Criterion for Informativeness

eTable 4 – Phase 4 Trials Not Meeting 4 Informativeness Criteria

eTable 5 – Trials Not Cited in Clinical Review Documents

### eMethods 1 – ClinicalTrials.gov search criteria

1. Condition or disease search terms:

a. ISCHEMIC HEART DISEASE: coronary artery disease OR coronary disease OR coronary heart disease OR coronary occlusion OR acute coronary syndrome OR myocardial ischemia OR angina pectoris OR angina, stable OR angina, unstable OR myocardial infarction OR ischemic heart disease
b. LUNG CANCER: Lung cancer OR lung neoplasm OR lung carcinoma OR non-small-cell lung cancer OR non-small-cell lung carcinoma OR small cell lung cancer OR small cell lung carcinoma OR lung tumor OR lung tumour
c. DIABETES: diabetes mellitus OR diabetes
2. Study type: “Interventional Studies (Clinical Trials)”
3. Recruitment status: “Recruiting, Completed, Suspended, Terminated, Active not recruiting, enrolling by invitation and unknown status”
4. Study start: 2009-01-01 to 2010-12-31

### eMethods 2 – Trial Inclusion and Exclusion criteria

*Inclusion criteria:*

- Primary outcome = clinical decision-related outcome: including mortality, morbidity, quality of life, functional status, need for further interventions, or an appropriate surrogate measures (for example, ejection fraction, Hgb A1c, or progression free survival in ischemic heart disease, diabetes mellitus and lung cancer respectively)
- Intervention of any type (drug, device, behavioral, surgical or other) directed towards the treatment or prevention of ischemic heart disease/lung cancer/diabetes mellitus (and not side-effects of the disease or complications from disease treatment)
- Trials with a US site
- Randomized trials
- Multi-arm trials
- Trials of interventions subject to FDA regulations and are FDA approved prior to trial start
- Trials of interventions not subject to FDA regulations
- Trials of interventions subject to FDA regulations, FDA approved post trial start, and have at least 5 years of follow-up since FDA approval, to allow for ample time for results incorporation into systematic reviews/clinical practice guidelines/UpToDate

*Exclusion criteria:*

- Exclusively evaluating safety, diagnostic or screening interventions
- Exclusion of phase 0 or phase 1 trials (given these early phase trials would be unlikely to inform clinical decision-making)
- Exclusion of extension studies with primary aim to enable continued access to drug (and without additional post-marketing surveillance outcomes)
- Indeterminate trials, which are ongoing trials have not surpassed double the allotted time for primary outcome completion (as first stated in the historical clinicaltrials.gov registration record)

### eMethods 3 – Flow Diagrams for Each Disease

Flow Diagram for Ischemic Heart Disease Interventional Trials

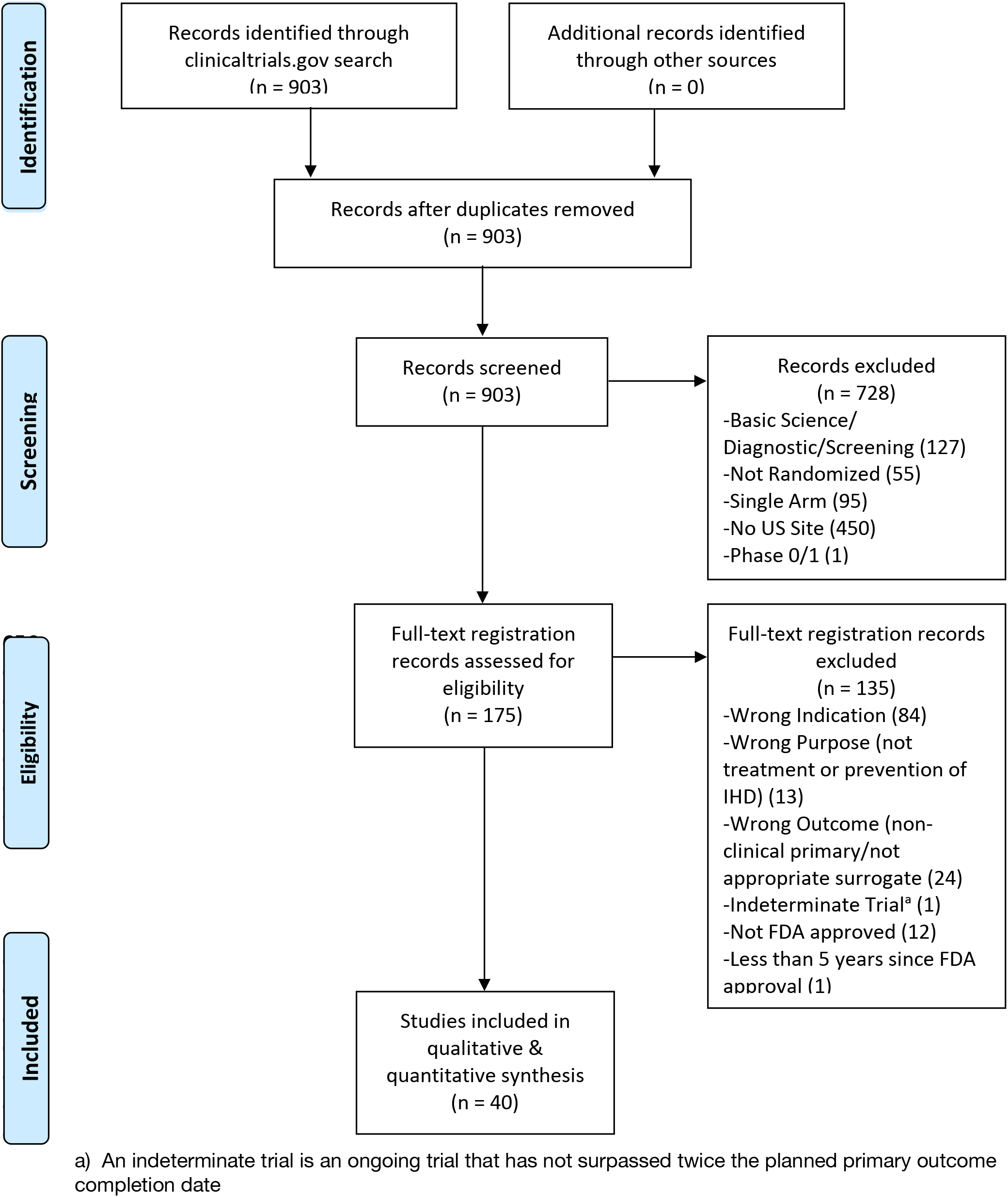

Flow Diagram for Diabetes Mellitus Interventional Trials

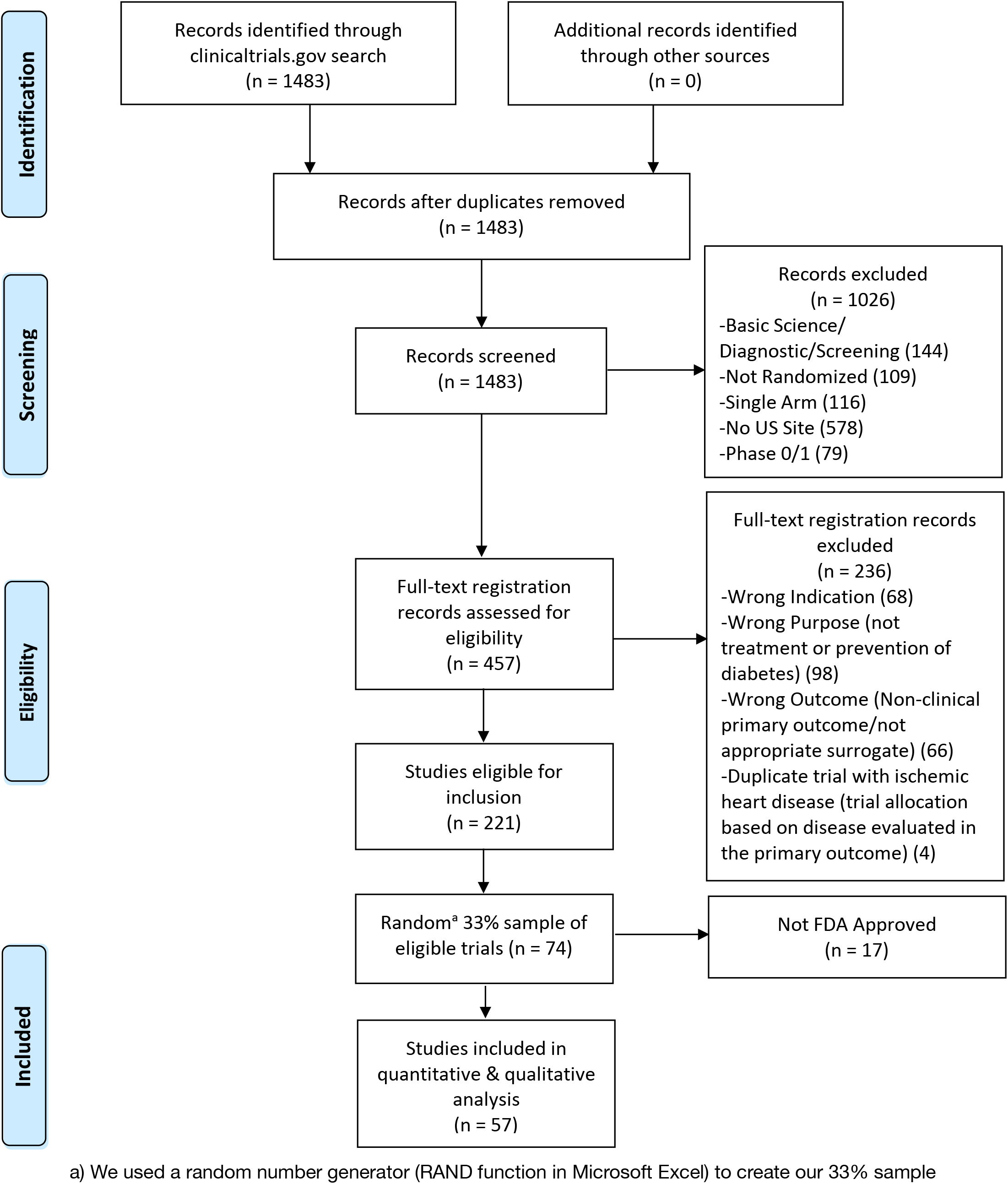

Flow Diagram for Lung Cancer Interventional Trials

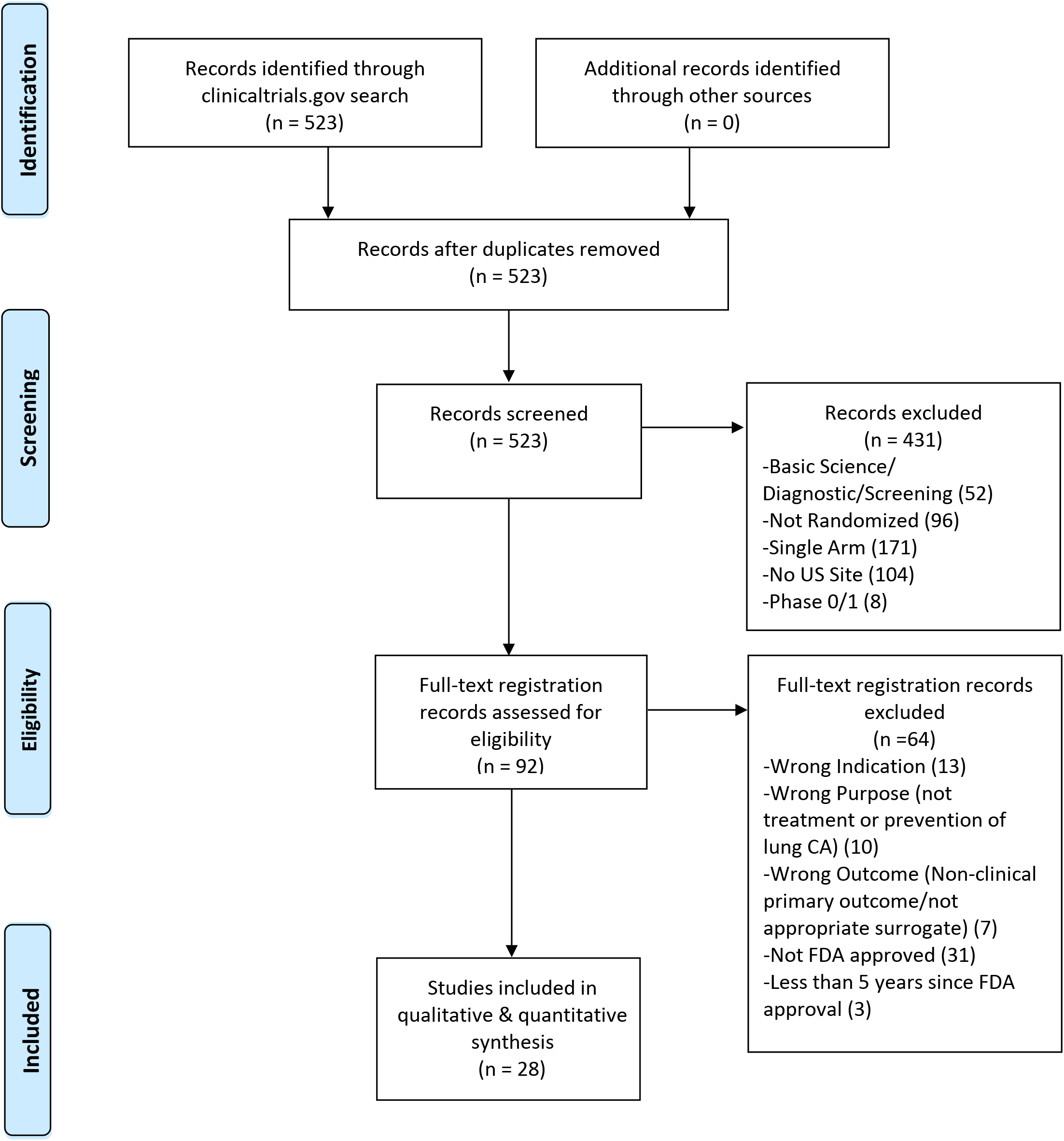

### eMethods 4 – Assessment of Regulatory Approval Status

Two authors (NH & HM) independently evaluated all eligible trials for regulatory approval status using Drugs@FDA^1^ for drug and biological interventions and the 510(k) Premarket Notification website for devices.^2^ Interventions were classified into one of 3 categories: i) FDA approved prior to trial start (drug, biological or device interventions approved for any use by the time of trial start); ii) FDA approved at least 5 years ago ((drug, biological or device interventions approved for any use prior to October 31, 2016); and, iii) interventions not subject to FDA approval.

### eTable 1 – Addressing the 4 Conditions for Informative Clinical Trials

| **Conditions for Informativeness<sup>1</sup>** | **Manner in which this is evaluated** |
| --- | --- |
| <u>Importance:</u><br>Trial hypothesis is likely to inform an important scientific, medical or policy question | -Trials are selected for inclusion in our cohort based on their potential to inform clinical decision-making, based on presence of a primary clinical outcome or appropriate surrogate.<br>-We also assess importance by evaluating the proportion of trials that are cited in clinical review documents (systematic reviews, clinical practice guidelines or point-of-care medical database articles). |
| <u>Design:</u><br>Trial methods are likely to provide meaningful evidence related to study hypothesis | -We assess trials that are designed to inform clinical decision-making for evidence of low risk of bias |
| <u>Feasibility:</u><br>Trial is likely to be feasible | -Feasible trials include:<br>-Completed trials that have reached $\geq 85\%$ planned recruitment<br>-Terminated trials stopped for an informative reason (such as efficacy, futility or safety)<br>-Ongoing trials that have not surpassed double their anticipated primary completion timeline |
| <u>Reporting:</u><br>Systems are in place to ensure timely, complete and accurate reporting | -A trial is reported if primary outcome results are made available through publication or results deposition on ClinicalTrials.gov |

<sup>1</sup>Column “Conditions for Informativeness” extracted from column1 in eTable 1<sup>3</sup>

### eMethods 5 – Flow Chart of Informative Criteria Asses

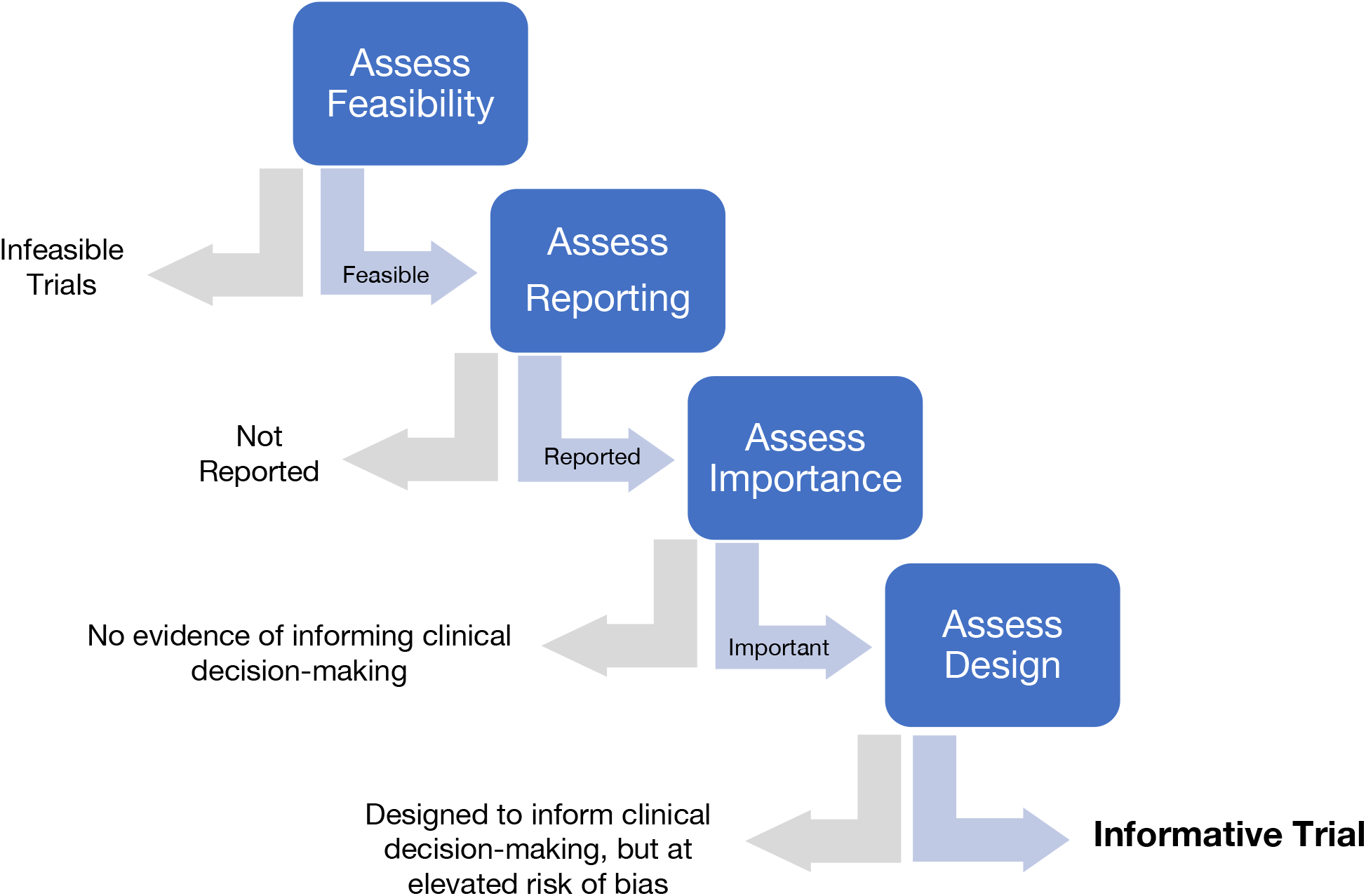

### eMethods 6 – Classification of Reason for Termination

| **NCT** | **Reason for Termination** | **Outcome** |
| --- | --- | --- |
| NCT00831441 | None Provided | Advanced to next step of assessment |
| NCT00863512 | None Provided | Advanced to next step of assessment |
| NCT00887315 | Accrual; Loss of sponsor | Infeasible |
| NCT00910299 | Futility | Advanced to next step of assessment |
| NCT00932152 | Accrual | Infeasible |
| NCT00965055 | Accrual | Infeasible |
| NCT01041781 | DSMB Recommendation | Advanced to next step of assessment |
| NCT01078272 | Accrual | Infeasible |
| NCT01246011 | Accrual | Infeasible |
| NCT00976677 | None Provided | Advanced to next step of assessment |
| NCT01177592 | Funding | Infeasible |
| NCT01179308 | PI closed study site | Infeasible |
| NCT01197963 | IRB Decision | Advanced to next step of assessment |
| NCT01231750 | Accrual | Infeasible |
| NCT01413750 | Accrual | Infeasible |

### eMethods 7 – Methodology for Publication Search

The search for publications was independently performed by two authors (NH & HM) and included an evaluation of publication links provided on ClinicalTrials.gov, as well as directed searches on Google Scholar, Scopus and Medline using a combination of the unique trial registration number (NCT number), surname of the principal investigator, indication, intervention, phase and study design. Publication identity was confirmed by comparing trial arms, sample size, intervention details, comparators and sponsor with the registration record. A published abstract was not counted as a full publication. Publication search was repeated in October 2021 by NH & HM for those trials without publications when first assessed.

### eMethods 8 – Systematic Review Citation Search Strategy and Quality Assessment

Assessment of citation of trial results in high quality systematic reviews (SRs) was independently performed by two authors (NH & HM). This first involved a search for sources that are well known for producing high quality SRs: Cochrane SRs on the Cochrane Database of Systematic Reviews^4^ and Agency for Healthcare Research and Quality (AHRQ) SRs ^5^. If trials were not included in Cochrane or AHRQ reviews, additional SRs for published studies were identified using the Scopus database ^6^ citation analysis search function or via Google Scholar ^7^ for unpublished studies. SRs identified through Scopus or Google Scholar that included trial results in review results were assessed for quality using a modified AMSTAR scoring system:

Operationalization of modified AMSTAR^8,9^ scoring system

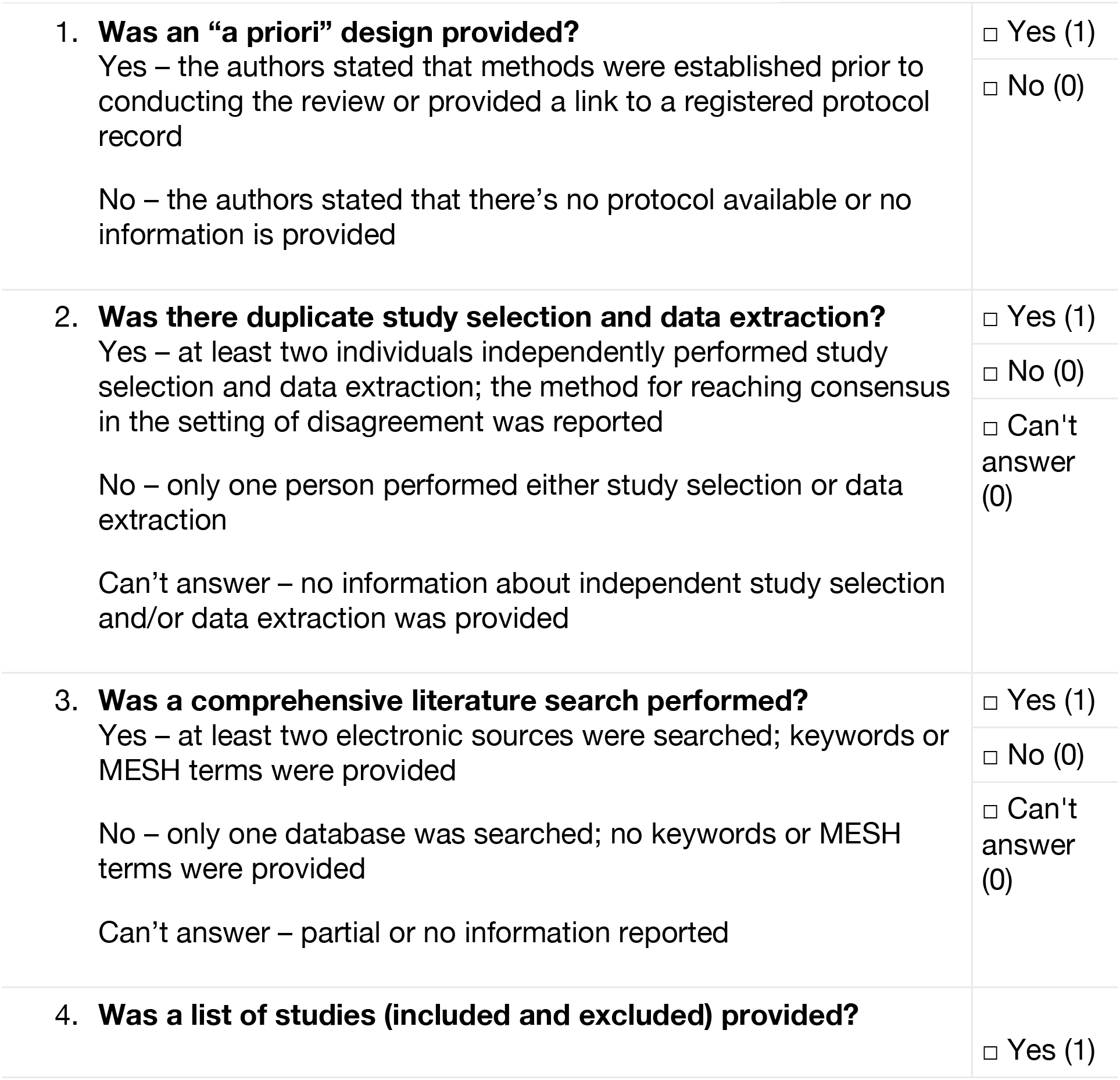
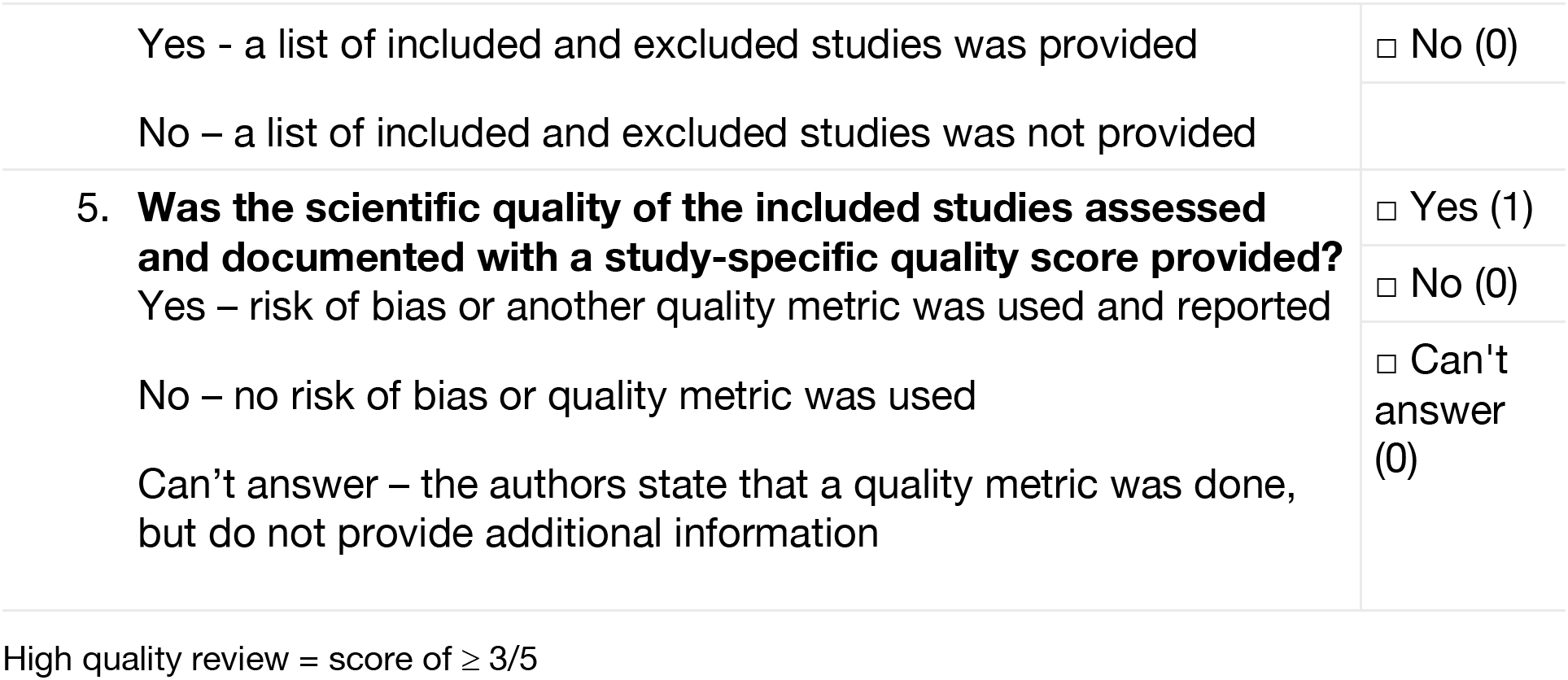

Trials were deemed to have fulfilled the importance criterion if they were cited in the results of a Cochrane SR, an AHRQ SR, or an SR achieving a modified AMSTAR score of greater or equal to 3 out of 5.

Assessment of citation of trial results in SRs was repeated in October 2021 by NH & HM for those trials without an informative citation when first assessed.

### eMethods 9 – Clinical Practice Guideline and Point-of-Care Medical Database Search Strategies and Quality Assessment

Assessment of citation of trial results in Clinical Practice Guidelines (CPGs) was independently performed by two authors (NH & HM). CPGs were identified via Scopus ^6^ citation analysis for published studies or via Google Scholar ^7^ for unpublished trials. Quality of CPGs was assessed using a modified AGREE II scoring system:

Operationalization of modified AGREE II ^10^ scoring system

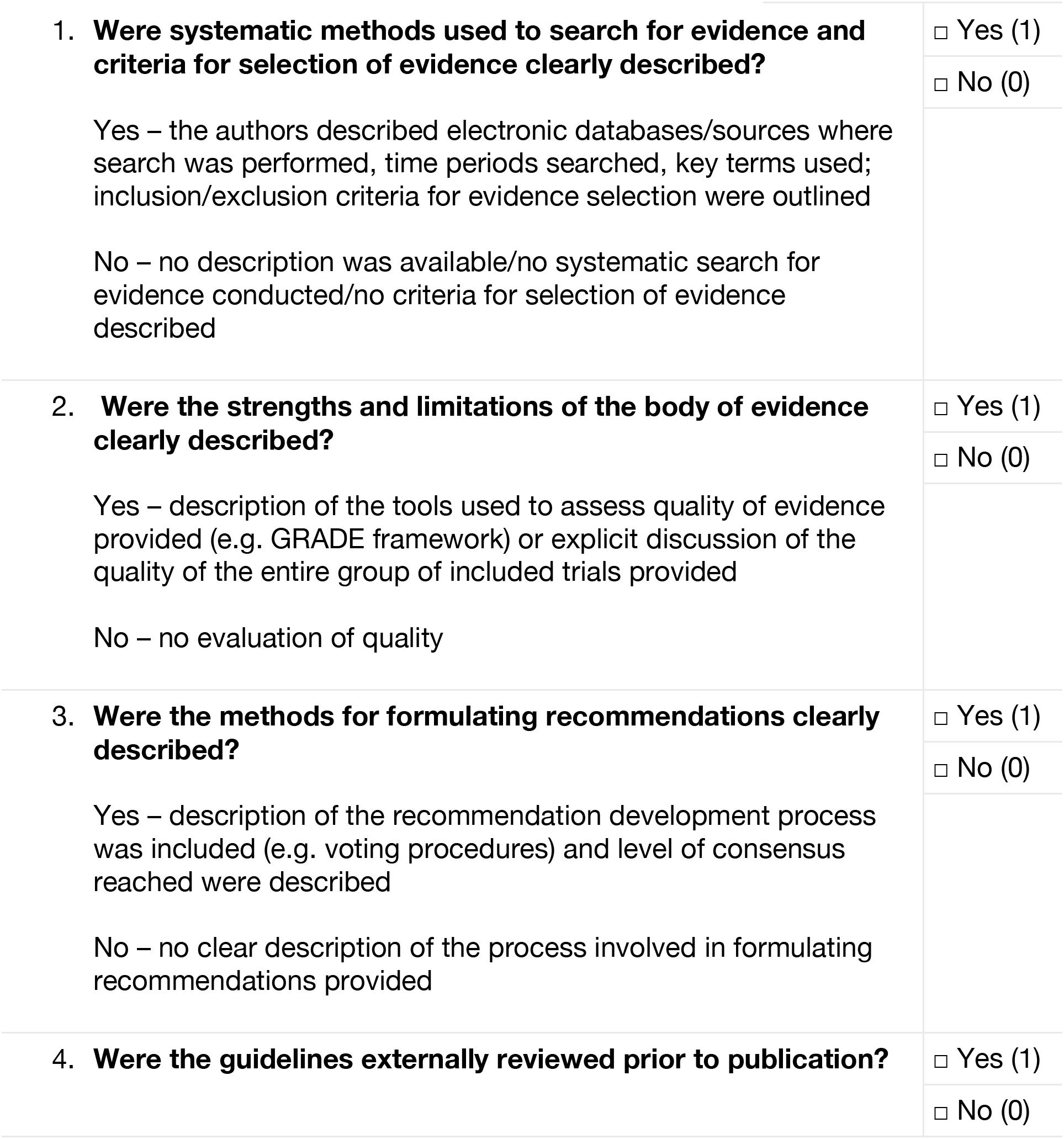
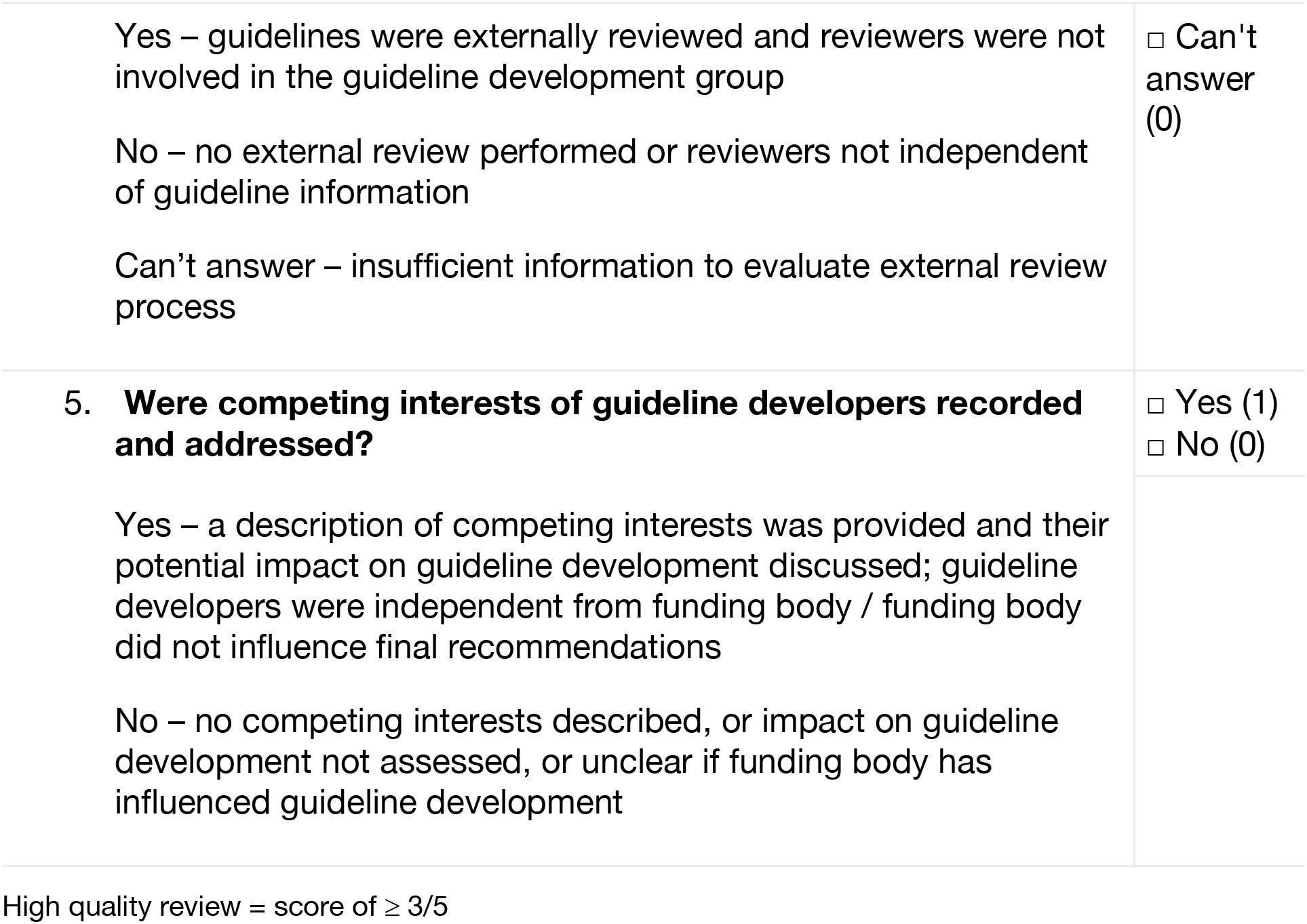

Trials were deemed to have fulfilled criteria for importance if they were cited in the results of a high-quality CPG. The remaining uncited trials were assessed for inclusion in a point-of-care medical database article by two authors (NH & HM). Using disease and intervention keywords, we searched UpToDate^11^ to identify any articles citing the remaining trials.

Assessment of citation of trial results in Clinical Practice Guidelines (CPGs) was repeated in October 2021 by NH & HM for those trials without an informative citation when first assessed.

### eMethods 10 – Operationalization of modified Cochrane Risk of Bias score

We employed a modified 2011 version of the Cochrane Risk of Bias Assessment Tool (detailed criteria for judging risk of bias provided in Table 8.5d of the Cochrane Handbook for Systematic Reviews of Interventions version 5.1) ^12^. When available, ROB scores were extracted directly from high-quality SRs identified during the assessment of trial importance. When not available, assessment was independently carried out by two authors (NH & HM), with differences resolved by a third (JK). Assessment included the following elements: i) random sequence generation; ii) allocation concealment; iii) blinding of participants and personnel; iv) blinding of outcome assessment; v) incomplete outcome data; and, vi) selective reporting. Trials were deemed of sufficient design quality if all elements were deemed to be “low risk of bias” or if a minority of elements were deemed of “unclear risk” and the remaining were “low risk.” Any “high risk” of bias element equated with poor trial design.

Of the 63 trials assessed for trial design, 36 ROB scores were extracted directly from SRs, the remaining 27 trials were assessed by the study team.

### eMethods 11 – Deviations to the Study Protocol

1. For our feasibility assessment, we first evaluated feasibility of goal patient enrollment and planned date of primary completion based on the first record available on ClinicalTrials.gov (this was independently double-coded). However, we repeated this assessment using the final registration record prior to trial start date as this was felt to provide a better evaluation of feasibility, allowing investigators to adjust enrollment plans and primary outcome timeline prior to trial start. The latter method was single coded and did not produce any change in the results of our feasibility assessment.
2. We performed two additional sensitivity analyses on the primary outcome: i) excluding all trials in the lower quartile of goal patient enrollment; and, ii) excluding all phase 1/2 and phase 2 trials from the assessment.
3. We excluded evaluation of primary outcome integrity from our assessment of trial informativeness, given concerns that there can be scientifically valid reasons for altering a primary outcome. For example, a primary outcome might be changed due to evolving clinical practice or in response to new data from outside the trial.
4. We excluded trials of interventions that were subject to FDA regulations, but were never approved for any indication, or were FDA approved after trial start, but did not have 5 years of follow-up time from 31 October 31 2016 post-approval to allow for enough time for trial results to be included in systematic reviews/clinical practice guidelines/UpToDate.
5. We excluded Indeterminate trials from our cohort, defined as ongoing trials that have not surpassed double the allotted time for primary outcome completion (as first stated in the historical clinicaltrials.gov registration record)

### eMethods 12 – STROBE Checklist for Cohort Studies

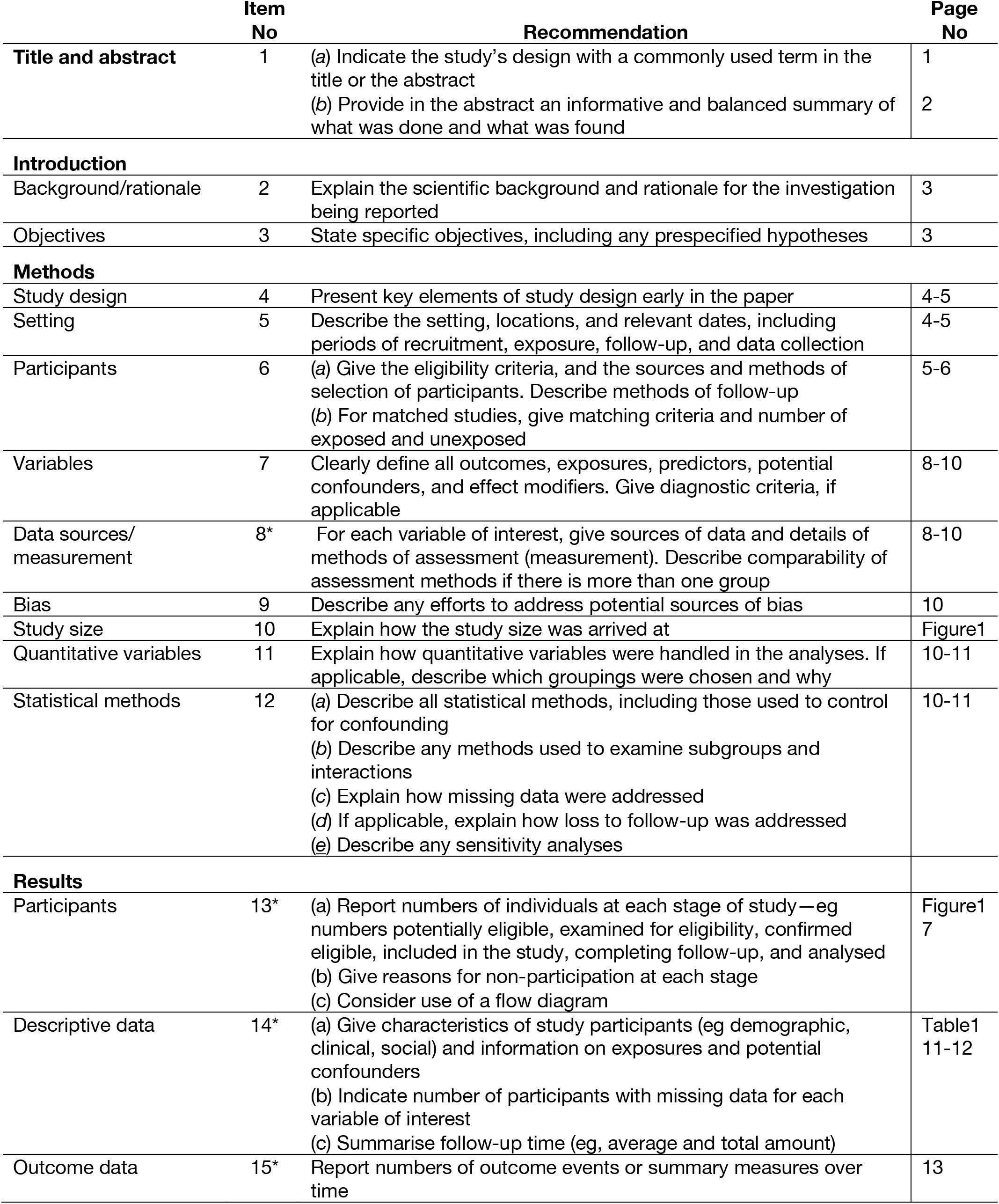
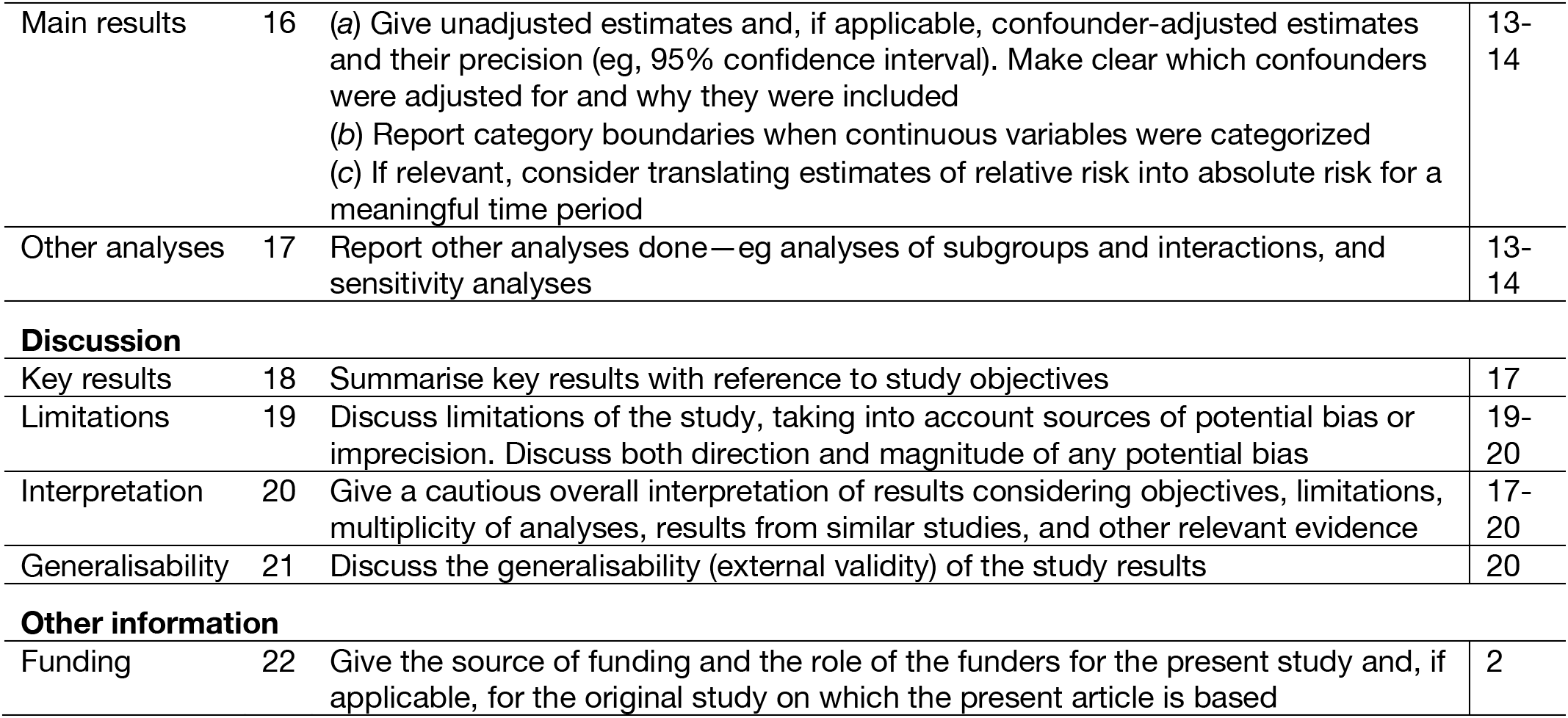

|  | Item No | Recommendation | Page No |
| --- | --- | --- | --- |
| **Title and abstract** | 1 | (a) Indicate the study's design with a commonly used term in the title or the abstract | 1 |
|  |  | (b) Provide in the abstract an informative and balanced summary of what was done and what was found | 2 |
| **Introduction** |  |  |  |
| Background/rationale | 2 | Explain the scientific background and rationale for the investigation being reported | 3 |
| Objectives | 3 | State specific objectives, including any prespecified hypotheses | 3 |
| **Methods** |  |  |  |
| Study design | 4 | Present key elements of study design early in the paper | 4-5 |
| Setting | 5 | Describe the setting, locations, and relevant dates, including periods of recruitment, exposure, follow-up, and data collection | 4-5 |
| Participants | 6 | (a) Give the eligibility criteria, and the sources and methods of selection of participants. Describe methods of follow-up<br>(b) For matched studies, give matching criteria and number of exposed and unexposed | 5-6 |
| Variables | 7 | Clearly define all outcomes, exposures, predictors, potential confounders, and effect modifiers. Give diagnostic criteria, if applicable | 8-10 |
| Data sources/<br>measurement | 8* | For each variable of interest, give sources of data and details of methods of assessment (measurement). Describe comparability of assessment methods if there is more than one group | 8-10 |
| Bias | 9 | Describe any efforts to address potential sources of bias | 10 |
| Study size | 10 | Explain how the study size was arrived at | Figure1 |
| Quantitative variables | 11 | Explain how quantitative variables were handled in the analyses. If applicable, describe which groupings were chosen and why | 10-11 |
| Statistical methods | 12 | (a) Describe all statistical methods, including those used to control for confounding<br>(b) Describe any methods used to examine subgroups and interactions<br>(c) Explain how missing data were addressed<br>(d) If applicable, explain how loss to follow-up was addressed<br>(e) Describe any sensitivity analyses | 10-11 |
| **Results** |  |  |  |
| Participants | 13* | (a) Report numbers of individuals at each stage of study—eg numbers potentially eligible, examined for eligibility, confirmed eligible, included in the study, completing follow-up, and analysed<br>(b) Give reasons for non-participation at each stage<br>(c) Consider use of a flow diagram | Figure1<br>7 |
| Descriptive data | 14* | (a) Give characteristics of study participants (eg demographic, clinical, social) and information on exposures and potential confounders<br>(b) Indicate number of participants with missing data for each variable of interest<br>(c) Summarise follow-up time (eg, average and total amount) | Table1<br>11-12 |
| Outcome data | 15* | Report numbers of outcome events or summary measures over time | 13 |
| Main results | 16 | (a) Give unadjusted estimates and, if applicable, confounder-adjusted estimates and their precision (eg, 95% confidence interval). Make clear which confounders were adjusted for and why they were included<br>(b) Report category boundaries when continuous variables were categorized<br>(c) If relevant, consider translating estimates of relative risk into absolute risk for a meaningful time period | 13-14 |
| Other analyses | 17 | Report other analyses done—eg analyses of subgroups and interactions, and sensitivity analyses | 13-14 |
| Key results | 18 | Summarise key results with reference to study objectives | 17 |
| Limitations | 19 | Discuss limitations of the study, taking into account sources of potential bias or imprecision. Discuss both direction and magnitude of any potential bias | 19-20 |
| Interpretation | 20 | Give a cautious overall interpretation of results considering objectives, limitations, multiplicity of analyses, results from similar studies, and other relevant evidence | 17-20 |
| Generalisability | 21 | Discuss the generalisability (external validity) of the study results | 20 |
| **Other information** |  |  |  |
| Funding | 22 | Give the source of funding and the role of the funders for the present study and, if applicable, for the original study on which the present article is based | 2 |

### eTable 2 – Inter-rater Agreement Rates

| **Category** | **Unweighted Cohen's Kappa** |
| --- | --- |
| Screening trials for inclusion/exclusion | 0.83 |
| Evaluating trial Feasibility | 0.98 |
| Evaluating trial Reporting | 0.79 |
| Evaluating trial Importance | 0.67 |
| Evaluating trial Design | 0.84 |

### eTable 3 – Proportion of Trials Meeting Each Criterion for Informativeness

| **Condition for Informativeness** | **Ratio** | **% (95% CI)** |
| --- | --- | --- |
| Feasibility | 90 of 125 trials | 72.0 (63.3 – 79.7) |
| Reporting | 81 of 90 trials | 90.0 (81.9 – 95.3) |
| Importance | 63 of 81 trials | 77.8 (67.2 – 86.3) |
| Design | 33 of 63 trials | 52.4 (39.4 – 65.1) |

### eTable 4 – Phase 4 Trials Not Meeting All 4 Informativeness Criteria

| **NCT** | **Disease** | **Title** | **Trial Status** | **Sponsor** | **Informativeness Assessment** |
| --- | --- | --- | --- | --- | --- |
| NCT00954707 | IHD | A Prospective, Randomized, Multi-Center, Double-Blind Trial to Assess the Effectiveness and Safety of Different Durations of Dual Anti-Platelet Therapy in Subjects Undergoing Percutaneous Coronary Intervention With the CYPHER® Sirolimus-eluting Coronary Stent (CYPHER® Stent) | Unknown (Previously, Active, NR) | Industry | Not cited in review documents |
| NCT00977938 | IHD | A Prospective, Multi-center, Randomized, Double-blind Trial to Assess the Effectiveness and Safety of 12 Versus 30 Months of Dual Antiplatelet Therapy in Subjects Undergoing Percutaneous Coronary Intervention With Either Drug-eluting Stent or Bare Metal Stent Placement for the Treatment of Coronary Artery Lesions | Completed | Non-Industry | Elevated Risk of Bias Score |
| NCT01050348 | IHD | A Double Blinded Randomized Placebo Controlled Study: To Investigate the Role of Upstream High Dose Statin Treatment in Patients With ST Segment Elevation Myocardial Infarction | Completed | Non-Industry | Not Reported |
| NCT01069003 | IHD | EDUCATE: a Prospective, Multi-center Study Designed to Collect Real-world Safety and Clinical Outcomes in Subjects Receiving One or More Endeavor Zotarolimus-Eluting Stents and Either Clopidogrel and Aspirin or Prasugrel and Aspirin as Part of a Dual Antiplatelet Therapy Drug Regimen | Completed | Industry | Elevated Risk of Bias Score |
| NCT01106534 | IHD | XIENCE V® Everolimus Eluting Coronary Stent System USA Post-Approval Study (XIENCE V® USA DAPT Cohort) (XVU-AV DAPT) | Completed | Industry | Poor Feasibility |
| NCT01178268 | IHD | XIENCE V Everolimus Eluting Coronary Stent System China: Post-Approval Randomized Control Trial | Completed | Industry | Not cited in review documents |
| NCT01221272 | IHD | A Phase 4, Randomized, Double-Blind, Placebo-Controlled, Cross-over Trial to Evaluate the Effects of Ranolazine on Myocardial Perfusion Assessed by Serial Quantitative Exercise SPECT Imaging | Completed | Industry | Poor Feasibility |
| NCT01230892 | IHD | The Evaluation of The Effects of Nebivolol in Comparison to Atenolol on Wall Shear Stress and Rupture Prone Coronary Artery Plaques in Patients With Moderate Coronary Artery Disease | Completed | Non-Industry | Not cited in review documents |
| NCT01246011 | IHD | Significance of Antibodies to Heparin/Platelet Factor 4 Complex in Vein Graft Patency and Potential Role of Argatroban for Prevention of Vein Graft Occlusion | Terminated | Non-Industry | Poor Feasibility |
| NCT01101867 | DM | Prandial Insulin Dosing Using the Carbohydrate Counting Technique in Hospitalized Patients With Diabetes | Completed | Non-Industry | Not cited in review documents |
| NCT01267448 | DM | A Pilot Study of Outpatient Discharge Therapy With Saxagliptin + Metformin XR or Sulphonylurea for Recently Diagnosed Type 2 Diabetes Presenting With Severe Hyperglycemia | Unknown (Previously, Recruiting) | Non-Industry | Poor Feasibility |
| NCT00978796 | DM | Pilot Study Assessing Glucose Effects of Sitagliptin (Januvia) in Adult Patients With Type 1 Diabetes | Completed | Non-Industry | Not cited in review documents |
| NCT00979628 | DM | Basal Bolus Versus Basal Insulin Regimen for the Treatment of Hospitalized Patients With Type 2 Diabetes Mellitus | Completed | Non-Industry | Elevated Risk of Bias Score |
| NCT01107717 | DM | Durability of Early Initial Combination Therapy With Exenatide/Pioglitazone/Metformin vs Conventional Therapy in New Onset Type 2 Diabetes | Active, NR | Non-Industry | Poor Feasibility |
| NCT00939250 | DM | A Comprehensive Intervention for Diabetes and Comorbid Depression in Primary Care | Completed | Non-Industry | Poor Feasibility |
| NCT00950677 | DM | The Effect of the Glucagon Suppressors Pramlintide and Exenatide on Postprandial Glucose Metabolism in Children With Type 2 Diabetes Mellitus | Completed | Non-Industry | Not Reported |
| NCT01221090 | DM | Employing Diabetes Self-Management Models to Reduce Health Disparities in Texas | Completed | Non-Industry | Elevated Risk of Bias Score |

IHD – Ischemic Heart Disease
DM – Diabetes Mellitus

### eTable 5 – Trials Not Cited in Clinical Review Documents

| **NCT** | **Disease** | **Title** | **Phase** | **Trial Status** | **Sponsor** | **Published Results** |
| --- | --- | --- | --- | --- | --- | --- |
| NCT00924118 | IHD | A Safety and Efficacy Evaluation of Sodium Nitrite Injection for the Prevention of Ischemia-Reperfusion Injury Associated with Acute Myocardial Infarction | 2 | Completed | Non-Industry | NO |
| NCT00954707 | IHD | A Prospective, Randomized, Multi-Center, Double-Blind Trial to Assess the Effectiveness and Safety of Different Durations of Dual Anti-Platelet Therapy (DAPT) in Subjects Undergoing Percutaneous Coronary Intervention with the CYPHER Sirolimus-eluting Coronary Stent | 4 | Unknown (Previously, Active, Not Recruiting) | Industry | NO |
| NCT01175018 | IHD | Anakinra to Prevent Adverse Post-infarction Remodeling | 2 | Completed | Non-Industry | YES |
| NCT01178268 | IHD | XIENCE V Everolimus Eluting Coronary Stent System China: Post-Approval Randomized Control Trial | 4 | Completed | Industry | NO |
| NCT01230892 | IHD | The Evaluation of The Effects of Nebivolol in Comparison to Atenolol on Wall Shear Stress and Rupture Prone Coronary Artery Plaques in Patients With Moderate Coronary Artery Disease | 4 | Completed | Non-Industry | YES |
| NCT01101867 | DM | Prandial Insulin Dosing Using the Carbohydrate Counting Technique in Hospitalized Patients With Diabetes | 4 | Completed | Non-Industry | YES |
| NCT01194245 | DM | A Phase II, Randomized, Double Blind, 2-Way Crossover Safety and Efficacy Study of Subcutaneously Injected Prandial Insulins: Lispro-PH20 or Aspart-PH20 Compared to Insulin Lispro (Humalog) in Patients With Type 1 Diabetes | 2 | Completed | Industry | NO |
| NCT00916357 | DM | Phase 2, Double-Blind Randomized, 3-way Cross-Over Liquid Meal Study With Optimal Doses of SC Administered Insulin Lispro With and Without rHuPH20 and Regular Human Insulin With rHuPH20 to Compare Pharmacokinetics, Postprandial Glycemic Response, and Optimal Insulin Dose in Patients With T2DM | 2 | Completed | Industry | YES |
| NCT00993824 | DM | Use of Continuous Glucose Monitoring With Ambulatory | 2/3 | Completed | Non-Industry | YES |
|  |  | Glucose Profile Analysis to Demonstrate the Glycemic Effect of Colesevelam HCl (Welchol) in Patients With Type 2 Diabetes |  |  |  |  |
| NCT00918138 | DM | A 4-Week, Multicenter, Randomized, Double-Blind, Phase 3b Trial to Evaluate the Efficacy of Saxagliptin in Combination With Metformin XR 1500 mg Versus Up-titrated Metformin XR to 2000 mg in Subjects With Type 2 Diabetes Who Have Inadequate Glycemic Control With Diet and Exercise and a Stable Dose of Metformin XR 1500 mg | 3 | Completed | Industry | YES |
| NCT00844090 | DM | The Role of Apathy in Glycemic Control | NA | Completed | Non-Industry | NO |
| NCT00978796 | DM | Pilot Study Assessing Glucose Effects of Sitagliptin (Januvia) in Adult Patients With Type 1 Diabetes | 4 | Completed | Non-Industry | YES |
| NCT01089569 | DM | Evaluation of Insulin Glargine and Exenatide: A Randomized Clinical Trial with Continuous Glucose Monitoring and Ambulatory Glucose Profile Analysis | NA | Completed | Non-Industry | YES |
| NCT00918203 | Lung CA | A Randomized Phase 2 Study of Human Anti-PDGFRα Monoclonal Antibody (IMC-3G3) with Paclitaxel/Carboplatin or Paclitaxel/Carboplatin Alone in Previously Untreated Patients with Locally Advanced or Metastatic Non-Small Cell Lung Cancer | 2 | Completed | Industry | YES |
| NCT00976677 | Lung CA | Randomized Double-Blind Placebo Controlled Phase II Trial Evaluating Erlotinib in Non-Smoking Patients With (Bevacizumab-Eligible and Ineligible) Advanced Non-Small Cell Lung Cancer | 2 | Terminated | Non-Industry | NO |
| NCT00977470 | Lung CA | Phase II Study of Erlotinib With or Without Hydroxychloroquine in Patients With Previously Untreated Advanced NSCLC and EGFR Mutations | 2 | Active, NR | Non-Industry | NO |
| NCT01085136 | Lung CA | Phase III Randomized Trial of BIBW 2992 Plus Weekly Paclitaxel Versus Investigator's Choice of Chemotherapy Following BIBW 2992 Monotherapy in Non-Small Cell Lung Cancer Patients Failing Previous Erlotinib or Gefitinib Treatment (LUX Lung 5) | 3 | Completed | Industry | YES |
| NCT01104155 | Lung<br>CA | Eribulin Mesylate in Combination<br>with Intermittent Erlotinib in<br>Patients with Previously Treated,<br>Advanced Non-Small Cell Lung<br>Cancer | 2 | Completed | Industry | YES |

IHD – Ischemic Heart Disease
DM – Diabetes Mellitus
Lung CA – Lung Cancer

